# A Bayesian Susceptible-Infectious-Hospitalized-Ventilated-Recovered Model to Predict Demand for COVID-19 Inpatient Care in a Large Healthcare System

**DOI:** 10.1101/2020.12.01.20241984

**Authors:** Stella Coker Watson Self, Rongjie Huang, Shrujan Amin, Joseph Ewing, Caroline Rudisill, Alexander C. McLain

**Author notes:** **Correspondence** Stella Self. Discovery Building, 901 Greene St, Columbia SC, 29208.

## Abstract

The COVID-19 pandemic strained healthcare systems in many parts of the United States. During the early months of the pandemic, there was substantial uncertainty about whether the large number of COVID-19 patients requiring hospitalization would exceed healthcare system capacity. This uncertainty created an urgent need for accurate predictions about the number of COVID-19 patients requiring inpatient and ventilator care at the local level. In this work, we develop a Bayesian Susceptible-Infectious-Hospitalized-Ventilated-Recovered (SIHVR) model to predict the burden of COVID-19 at the healthcare system level. The Bayesian SIHVR model provides daily estimates of the number of new COVID-19 patients admitted to inpatient care, the total number of non-ventilated COVID-19 inpatients, and the total number of ventilated COVID-19 patients at the healthcare system level. The model also incorporates county-level data on the number of reported COVID-19 cases, and county-level social distancing metrics, making it locally customizable. The uncertainty in model predictions is quantified with 95% credible intervals. The Bayesian SIHVR model is validated with an extensive simulation study, and then applied to data from two regional healthcare systems in South Carolina. This model can be adapted for other healthcare systems to estimate local resource needs.

## 1 INTRODUCTION

The World Health Organization declared the COVID-19 outbreak a global pandemic on March 11th, 2020. COVID-19 is a respiratory disease caused by the SARS-CoV-2 virus, which spreads primarily from person to person through respiratory droplets ^1^. The pandemic spread rapidly across the globe, with over 34,287,000 infections and 1,023,000 deaths reported worldwide by October 1st, 2020^2^. The United States (US) accounts for approximately 21.2% and 20.2% of world cases and deaths, respectively ^2^. Approximately 14% individuals with COVID-19 require hospitalization ^3^. In late winter and early spring of 2020, the rapid spread of COVID-19 threatened to overwhelm healthcare systems with more patients than the systems could accommodate. In the US, these concerns caused state governments to close nonessential businesses, issue stay at home orders, and mandate other forms of social distancing in an attempt to mitigate the spread of the virus. While such efforts initially prevented large outbreaks in many states, the pandemic nevertheless strained healthcare systems in some parts of the country, most notably in New York City. The gradual easing of these restrictions during the late spring and summer of 2020 caused a resurgence of COVID-19 in many states, creating substantial uncertainty about the trajectory of the pandemic in these areas. The post-reopening resurgence strained inpatient and intensive care unit (ICU) capacity in several areas of the US, most notably in Texas and Arizona. With policymakers under increasing pressure to avoid reimposing lockdowns and similar restrictions, the capacity of the healthcare system to treat COVID-19 patients is an essential metric for making policy decisions.

It is critically important to accurately predict local demand for COVID-19-related care. Such predictions provide advance notice if the demand for inpatient or ventilator care is likely to exceed the available capacity. This advanced warning gives healthcare systems time to expand capacity by increasing staffing, purchasing more ventilators, converting regular beds to ICU beds, reducing elective procedures to leave more resources for COVID-19 patients, and, in extreme cases, constructing temporary field hospitals. Even when demand for COVID-19 inpatient care is unlikely to exceed the available capacity, as is currently the case for much of the US, such predictions are still valuable for making staffing decisions, determining the ideal capacity for designated ‘COVID-19 wards’, and estimating the amount of personal protective equipment needed. Furthermore, appropriate uncertainty intervals are necessary to ensure that healthcare systems are prepared for worst case scenarios.

A variety of models exist for predicting the burden of COVID-19. The Institute for Health Metrics and Evaluation (IHME) model predicts the number of COVID-19 deaths using a curve fitting technique, and then uses the estimated number of deaths to estimate hospital resource use ^4^. While the IHME model provides state level estimates for resource use, it does not provide estimates on the health system level, limiting its usefulness for local health system planning ^4^. The University of Massachusetts Amherst model uses of a number of other models to create an ensemble model, which is robust to small fluctuations in observed data, but is again not customizable to the health system level ^5^. Numerous other models exist for predicting COVID-19 cases, hospitalizations, ventilations, and mortality. The Centers for Disease Control and Prevention (CDC) provides a helpful summary of existing prediction models on their webpage ^6^.

Among the many models for predicting the burden of COVID-19 are a number of compartmental models such as susceptible-infectious-recovered (SIR) and susceptible-exposed-infectious-recovered (SEIR) models. Some of these models are deterministic and estimate key parameters from data prior to fitting the model. Others attempt to find optimal parameters values by fitting model projections to observed data and quantifying the resulting uncertainty. The COVID-19 Hospital Impact Model for Epidemics (CHIME) is a deterministic SIR model which is customizable to the health system level ^7^. However, the CHIME model quantifies uncertainty in predictions via a sensitivity analysis which requires many assumptions about the relative likelihood of various parameter values rather than estimating the uncertainty directly from the observed data. Pei and Shaman (2020) use a deterministic metropolitan SEIR model which accounts for the movement of people between counties to forecast county-level COVID-19 incidence for the US ^8^. The Differential Equations Leads to Predictions of Hospitalizations and Infections (DELPHI) model from the COIVDAnalytics group at the Massachusetts Institute of Technology is another deterministic SEIR model, with additional model states to account for undocumented infections, hospitalizations, and individuals in quarantine ^9^. A major drawback of deterministic SIR and SEIR approaches is that they fail to appropriately quantify the uncertainty in their estimates. Quantifying the uncertainty in model projections of the utmost importance, as the trajectory of the COVID-19 pandemic is highly dynamic and sensitive to changes in social distancing, masking, and other societal dynamics. To make the best decisions and most appropriate contingency plans, healthcare systems need to quantify the relative likelihood of a range of possible scenarios, rather than basing decisions on a single point estimate of the number of individuals requiring care. One way to improve the uncertainty quantification of compartmental models is to embed a SIR or SEIR system of differential equations inside a Bayesian statistical model. This approach allows one to combine the predictive capabilities of the SIR/SEIR model with the tractable inference framework of the Bayesian paradigm. De Brouwer et al. (2020) use a Bayesian SIER model to estimate the effectiveness of various COVID-19 containment measures implemented in different counties ^10^. Hidaka and Torii (2020) use a Bayesian SIR model to predict the number of COVID-19 cases in the US and several other countries ^11^. However, neither of these models allow for local hospital resource use estimation.

In this paper, we present a Bayesian susceptible-infectious-hospitalized-ventilated-recovered (SIHVR) model; this model is similar to the standard SIR model, but includes additional states for hospitalization and ventilation. Our model is designed to make predictions at the healthcare system level. Our model predicts the daily number of reported COVID-19 cases, the daily number of COVID-19 inpatient admissions, daily census of hospitalized non-ventilated of COVID-19 patients, and daily census of ventilated COVID-19 patients. These quantities are assumed to follow a negative binomial distribution whose mean is governed by the solution to the SIHVR system of differential equations. The Bayesian framework allows us to estimate model parameters from the observed data and quantify the uncertainty in our parameter estimates. To our knowledge, our model is the only existing Bayesian SIHVR model for predicting the burden of COVID-19 at the healthcare system level that includes time varying transmission and hospitalization rates, and simultaneously utilizes hospital admissions data, inpatient census data, and ventilated census data, as well as the number of reported COVID-19 cases and social distancing metrics at the county level. We describe our model in Section 2, demonstrate its performance via an extensive simulation study in Section 3, and evaluate its real-world predictive performance using data from the Prisma Health System in South Carolina in Section 4. Section 5 provides concluding remarks.

## 2 METHODOLOGY

### 2.1 The Data

Our Bayesian SIHVR model integrates data from the Prisma Health System, the South Carolina Department of Health and Environmental Control (SCDHEC), and Unacast’s social distancing metrics (derived from mobile phone GPS location data). The Prisma Health System includes two major regional systems serving patient populations in geographically distinct parts of South Carolina, the Upstate System and the Midlands System. The daily number of SARS-CoV-2 positive admissions to each regional system was obtained from March 6th, 2020 (the date of the first reported case of COVID-19 in South Carolina) to July 15th, 2020. The total number of SARS-CoV-2 positive individuals in inpatient care and not on a ventilator (non-ventilated census) and total number of SARS-CoV-2 positive individuals on a ventilator (ventilated census) was obtained for each system over the same time period.

The number of confirmed new SARS-CoV-2 infections reported each day was obtained from SCDHEC. These cases are aggregated to the county level, and publicly available ^12^. A confirmed case is defined as an individual who had a positive polymerase chain reaction or antigen test for SARS-CoV-2 conducted via nose or throat swab, regardless of whether the individual had symptoms of infection or not. The reporting date reflects the date the case was publicly announced by SCDHEC.

Social distancing data was obtained from the Unacast Social Distancing Scoreboard (USDS). Specifically, we used the ‘Change in Non-Essential Visits’ metric, which estimates the daily percent change in visits to non-essential businesses over pre-pandemic levels (defined as the 4 week period prior to March 8th, 2020) by tracking GPS locations from mobile phones ^13^. Not all SARS-CoV-2 infections are documented by a positive viral test for SARS-CoV-2. Evidence from SARS-CoV-2 antibody tests suggests there are 6-24 unreported infections for each reported infection ^14^. Reliable data on antibody seroprevalence over time are currently not available for South Carolina. As a result, we use the daily number of confirmed COVID-19 cases reported each day by SCDHEC. The bias in the number of confirmed versus actual cases has led to the development of variants of the SIR model (e.g. Hao et al., 2020^15^), which aim to estimate the number of actual cases over time using assumptions about dynamics of COVID-19 transmission. Here, our goal is to predict the number of SARS-CoV-2 positive patients requiring inpatient and ventilator care, not the actual number of SARS-CoV-2 positive individuals. As a result, the bias in the number of confirmed infections is not an issue as long as we can relate the number of confirmed cases to the number of cases requiring inpatient and ventilator care.

Another issue is that the available data consists of the number of confirmed or suspected COVID-19 cases requiring inpatient or ventilator care, which may differ from the true number of such cases. Fortunately, most COVID-19 cases requiring hospitalization are documented with a positive SARS-CoV-2 viral test, as testing anyone presenting with COVID-19-like symptoms for active SARS-CoV-2 infection has become standard practice. As a result, almost all hospitalized cases of COVID-19 are documented with a positive test result, and under-detection of hospitalized cases is not of major concern. We examine the effects of under-detection in case incidence via simulation in study in Section 3, and find that while under-detection does impede our model’s ability to accurately estimate the number of cases in a given county, its effects on the model’s ability to estimate the number of patients hospitalized and ventilated (of primary interest here) is minimal.

### 2.2 The Data Likelihood

Let *U*_*ct*_ denote the number of COVID-19 cases reported from area *c* on day *t*, for *c* = 1, 2, …, *C*, and *t* = 1, 2, …, *T*, where *C* is the number of areas (e.g. counties, census tracts, etc.) served by the healthcare system in question, and *T* is the number of days of data we wish to include in the model. Furthermore, let *W*_*t*_ denote the number of COVID-19 patients admitted to inpatient care on day *t, Y*_*t*_ denote the total number of hospitalized, non-ventilated patients on day *t*, and *Z*_*t*_ denote the total number of ventilated patients on day *t*. We assume:

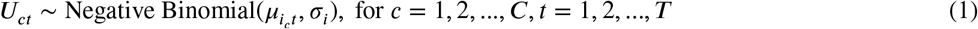

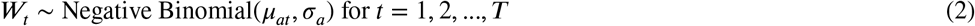

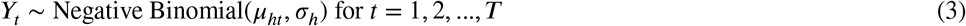

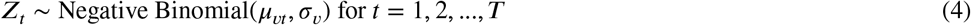

where *X* ∼ Negative Binomial(*µ, σ*) indicates that the random variable *X* follows a negative binomial distribution with mean *µ* and size σ, where 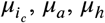, and *µ*_*v*_ are determined by the SIHVR system of differential equations specified in detail below and σ_*i*_ σ_*a*_, σ_*h*_ and σ_*v*_ are non-negative. Furthermore, we assume 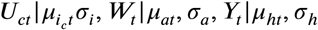 and *Z*_*t*_ |*µ*_*vt*_, σ_*v*_ are independent for all *c*, = 1,…, *C* and *t* = 1, …, *T*.

### 2.3 The SIHVR Model

We assume the population of each area is composed of susceptible, infectious (but not hospitalized), hospitalized (but not ventilated), ventilated, and recovered individuals. Let *N*_*c*_ denote the population of area *c*, and let *S*_*c*_(*t*), *I*_*c*_(*t*), *H*_*c*_(*t*), *V*_*c*_(*t*), and *R*_*c*_(*t*) denote the number of susceptible individuals, infectious non-hospitalized individuals, hospitalized non-ventilated individuals, ventilated individuals, and recovered individuals from area *c* at time *t*, respectively. As we are not interested in predicting the number of COVID-19 deaths, we treat dead and recovered patients as single category (recovered); as our model assumes recovered patients are no longer susceptible to infection, incorporating the dead patients into the recovered state has little effect on model dynamics.

We assume that the number of individuals in each state at time *t* is governed by the solution to the following system of differential equations:

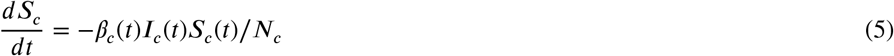

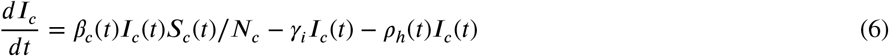

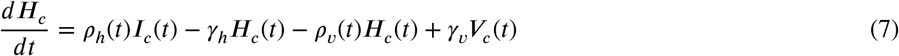

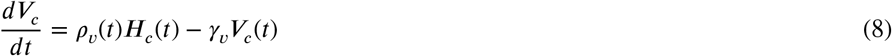

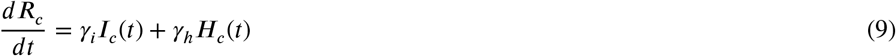

for *c* = 1, …, *C* and *t* ∈ [0, *T*]. Here, *γ*_*i*_ ∈ (0, 1) is the recovery rate for non-hospitalized individuals, *γ*_*h*_ ∈ (0, 1) is the discharge rate of non-ventilated individuals, and *γ*_*v*_∈ (0,1) is the rate at which ventilated individuals are removed from the ventilator. The transmission rate for area *c* (*β*_*c*_ (*t*)), the proportion of infected people entering the hospital each day *ρ*_*h*_(*t*)), and the proportion of hospitalized patients beginning ventilation each day (*ρ*_*v*_ (*t*)) are time-varying. We assume they have the following form:

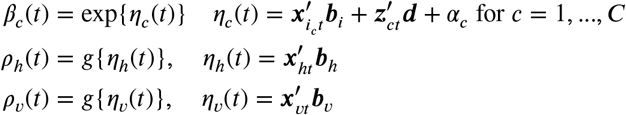

where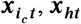, and ***x***_***vt***_ are B-spline basis functions in time evaluated at *t, **b***_*i*_,***b***_*h*_ and ***b***_*v*_ are the associated vectors of coefficients, ***z***_***ct***_ is a vector of social distancing metrics from area *c* associated with day *t*, ***d*** are the associated coefficients, α_*c*_ is a random effect for area *c* and *g*(·) is the inverse logistic function. For more on B-splines, see De Boor (1978) ^16^. Figure 1 provides an illustration of the various model compartments and the flow of individuals between them. For more on SIR and other compartmental models, see Brauer et al. (2008) ^17^ or Tolles and Luong (2020) ^18^.

**FIGURE 1.**
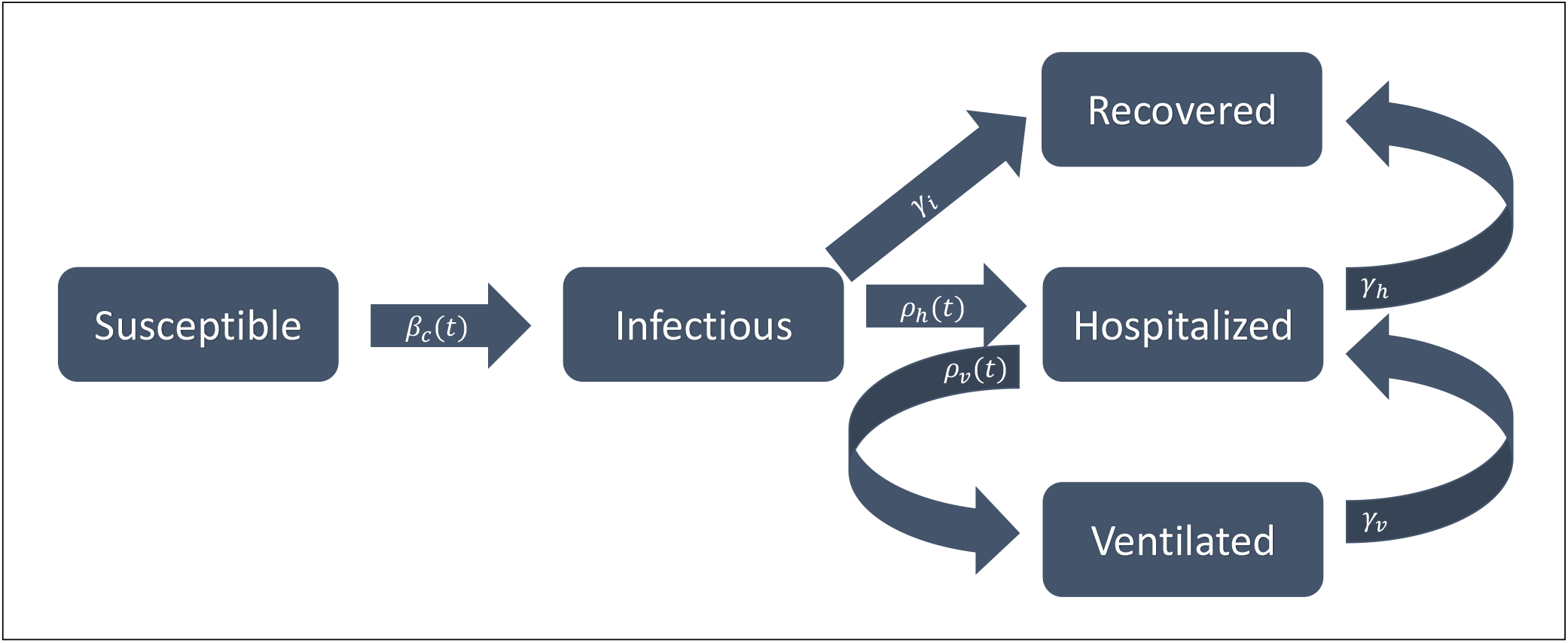
An illustration of the SIHVR model. Rectangles represent compartments, and arrows indicate the flow of individuals between compartments.

The above model results in 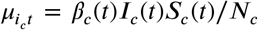, that is, the mean number of new confirmed SARS-CoV-2 infections in area *c* on day *t* is equal to the rate of new infections in area *c* on day *t*. We also assume 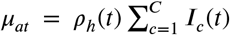, that is, the mean number of SARS-CoV-2 positive individuals admitted to inpatient care on day *t* is given by rate of flow into the hospitalization state on day *t*, summed over all areas. Finally, we assume 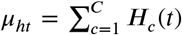, that is, the mean number of non-ventilated individuals in inpatient care on day *t* is equal to the total number of indiduals in the non-ventilated state on day *t*, and 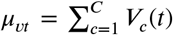, the mean number of ventilated individuals on day *t* is equal to the total number of individuals in the ventilated state at time *t*.

## 2.4 Initial Conditions

It is also necessary to specify the initial conditions for the SIHVR system of differential equations. Limited testing during the early phases of the pandemic makes it likely that the initial number of infections in each area was non-zero prior to the first reported case. To account for this, we treat *I*_*c*_(0) as unknown, and estimate it along with the other parameters. We assume that the initial number of people in the hospitalized, non-ventilated state, ventilated state and recovered state are known constants, and that all individuals who were not initially infectious, hospitalized, ventilated, or recovered were susceptible.

### 2.5 Prior Distributions

To fully specify our Bayesian model, it is necessary to assign prior distributions to all unknown parameters. We assume the following weakly informative prior distributions:

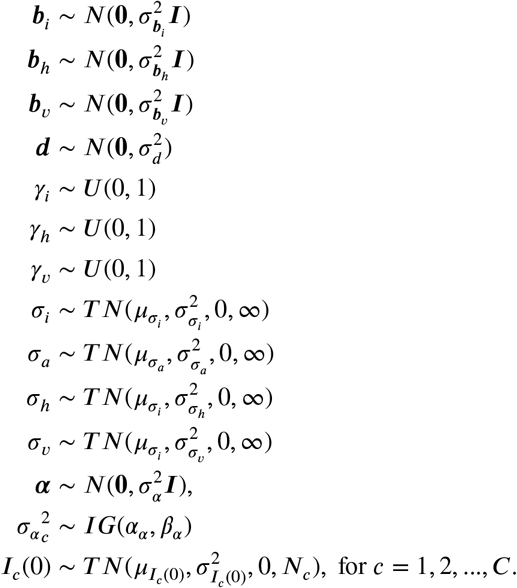

Here ***X*** ∼ *N*(***µ, A***) denotes that the random vector *X* follows a multivariate normal distribution with mean vector ***µ*** and variance-covariance matirx ***A***, *X* ∼ *U* (*a, b*) denotes that the random variable *X* follows a uniform distribution on the interval (*a, b*), *X* ∼ *T N*(*µ*, σ^2^,*a, b*) denotes that the random variable *X* follows a truncated normal distribution supported on the interval (*a, b*) with mean *µ*, and variance σ^2^, *X* ∼ *IG*(*a, b*) denotes that the random variable *X* follows an inverse gamma distribution with shape parameter and scale parameter *b*, 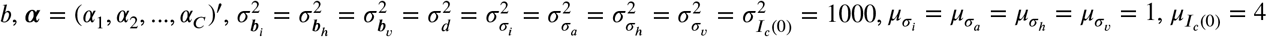, and *α*_*α*_ = *β*_*α*_ = 2.

### 2.6 Model Fitting Procedure

We developed a Markov chain Monte Carlo (MCMC) algorithm to generate a sample of the unknown parameters from the posterior distribution. As the full conditional distributions of most model parameters are not recognizable, the sampling routine consists of Metropolis Hasting steps. For the full sampling algorithm, see Web Appendix A. After generating a posterior parameter sample of size *G*, the SIHVR system of differential equations was solved *G* times, once for each set of sampled parameters, thereby generating a posterior sample of 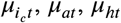, and *µ*_*vt*_, for *c* = 1, 2, …, *C* and *t* = 1, 2, …, *T*. As the distribution of these parameters was right skewed, the posterior median was used as point estimate, rather than the posterior mean. This sample, along with the posterior sample of *σ*_*i*_, *σ*_*a*_, *σ*_*h*_, and *σ*_*v*_ was used to generate a sample from the posterior predictive distribution of *U*_*ct*_, *W*_*t*_, *Y*_*t*_ and *Z*_*t*_, for *c* = 1, 2, …, *C* and *t* = 1, 2, …*T*. Specifically, for each value of *c* and *t* and for each pair 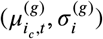 in the posterior sample, 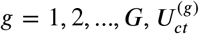 was generated from a negative binomial distribution with mean 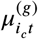 and size 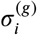. Posterior samples for the *W*_*t*_s, *Y*_*t*_s and *Z*_*t*_s were generated analogously using the appropriate distributions. These samples were then used to create posterior prediction intervals for each quantity in the usual way. The MCMC algorithm was run for 20,000 iterations, with the first 10,000 iterations discharged as the burn in period. Convergence was assessed via trace plots.

## 3 SIMULATION STUDY

In this section, we evaluate the performance of the Bayesian SIHVR model with a simulation study. We first consider the case of perfect case detection, i.e. we assume all COVID-19 cases are documented with a positive test result. We then consider the effect of under-detection on our methodology’s performance. For each of these scenarios, we evaluate ‘early phase’ performance using *T* = 57 days of data (the equivalent of using data from the first SCDHEC reported case on March 6th to May 1st) and ‘later phase’ performance, using *T* = 118 days of data (the equivalent of using data from March 6th to July 1st). For each dataset, the data from days *t* = 1, …, *T* was used to fit the model, and data from days *t* = *T* + 1, …, *T* + 14 was used to assess out of sample prediction performance.

To generate data, we take *C* = 2 areas, and solve the system of differential equations given by (5)-(9). The transmission rate and hospitalization rate are chosen so that the observed number of cases and hospitalizations are similar to the observed data from the Prisma Health Upstate System during time period under consideration (March 6th to May 1st for the early stage and March 6th to July 1st for the late stage). As a result, these parameter specifications vary somewhat across the 4 simulation studies (for the specific conditions used for each of the 4 studies see Web Appendix B). To mimic the noisy behavior of the social distancing metric, the observed USDS metric from Greenville and Spartanburg counties in the Upstate region of South Carolina in was used as the social distancing metric in the transmission rate for data generation. A piecewise cubic B-spline basis with 4 (3) equally spaced knots was used to estimate the transmission (hospitalization) rate. We took ***z***_*ct*_ = *z*_*ct*−14_, the social distancing metric observed in county *c* on day *t* − 14, *ρ*_*v*_ (*t*) = 0.05, *γ*_*i*_ = 1/14, *γ*_*h*_= 1/10, *γ*_*v*_= 1/10, *N*_1_ = 498402 and *N*_2_ = 302195 (the populations of Greenville and Spartanburg counties). After solving the system of differential equations given in (5)-(9), *U*_*ct*_, *W*_*t*_, *Y*_*t*_, and *Z*_*t*_, for *c* = 1, …, *C*, and *t* = 1, …, *T* + 14 were independently generated from Poisson distributions with means given by 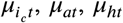 and *µ*_*vt*_, respectively. For *T* ∈ {57, 118}, 500 independent datasets were generated as described above. To assess effect of under-detection on the method, we generated 500 more datasets for each value of *T*, this time generating *U*_*ct*_ from a Poisson distribution with mean 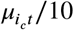, that is, we assume only 10% of cases are detected on average.

Tables 1 and 2 and Figure 2 summarize the results of our study. Specifically, Table 1 summarizes the results for the non-ventilated census, ventilated census, and hospital admissions. For each quantity, the table provides the estimated mean empirical bias, the mean absolute out of sample prediction error, the mean absolute out of sample prediction percent error, defined as the absolute bias divided by the true value multiplied 100, and the empirical coverage probability of 95% prediction intervals based on out of sample data. The empirical bias is averaged over days *t* = 1, …, *T*, and the other quantities are averaged over days *t* = *T* +1, …, *T* +14. Web Table 1 in Web Appendix C provides similar information for the area-level reported case incidence. Figure 2 provides plots of the posterior median estimates, true values used for data generation, and 95% credible prediction intervals for the non-ventilated census, ventilated census, and new admissions over time, averaged over all 500 datasets. Web Figure 2 in Web Appendix C displays similar results for the estimated area-level reported case incidence and area-level transmission rates. Note that as our primary goal is providing the healthcare system with a range of patient numbers requiring non-ventilated and ventilated inpatient care, the credible intervals are 95% credible intervals for *prediction*. Thus, we would expect roughly 95% of observations to fall within the interval, rather than expecting the interval to capture the true parameter value 95% of the time. Table 2 summarizes the results for key model parameters (*γ*_*h*,_ *γ*_*v*_), providing the posterior mean estimate, and associated empirical bias, empirical MSE, empirical standard deviation, and empirical coverage probability (ECP) for 95% credible intervals. Web Table 2 in Web Appendix C provides the same information for *γ*_*i*._

**TABLE 1.** Summary of Simulation Study Results: The table provides the empirical bias (averaged over days 1, 2, …, *T* and the 500 datasets), empirical absolute prediction error (averaged over days *T* + 1, *T* + 2, …, *T* + 14 and 500 datasets), empirical absolute prediction percent error (averaged over days *T* + 1, *T* + 2, …, *T* + 14 and the 500 datasets), and empirical coverage probability for 95% prediction intervals (averaged averaged over days *T* + 1, *T* + 2, …, *T* + 14 and the 500 datasets).

| Quantity | % Reported | Bias | Abs. Pred. Er. | % Abs. Pred. Er. | 95% ECP |
| --- | --- | --- | --- | --- | --- |
| *Early Phase (March 6 to May 1), $T = 57$* |  |  |  |  |  |
| Non-Ventilated Census | 100% | 0.0026 | 11.5243 | 12.38 | 0.9966 |
|  | 10% | 0.0121 | 5.7292 | 15.42 | 0.9936 |
| Ventilated Census | 100% | 0.0124 | 3.0082 | 11.21 | 0.9940 |
|  | 10% | 0.0156 | 1.6354 | 13.22 | 0.9861 |
| Admissions | 100% | 0.0107 | 2.2893 | 14.63 | 0.9917 |
|  | 10% | -0.0013 | 1.1241 | 21.12 | 0.9836 |
| *Later Phase (March 6 to July 1), $T = 118$* |  |  |  |  |  |
| Non-Ventilated Census | 100% | 0.0455 | 3.1391 | 4.77 | 0.9974 |
|  | 10% | 0.0908 | 2.9144 | 7.73 | 0.9943 |
| Ventilated Census | 100% | -0.0088 | 1.3656 | 3.94 | 0.9941 |
|  | 10% | 0.0337 | 1.0460 | 4.89 | 0.9901 |
| Admissions | 100% | -0.0072 | 0.4573 | 8.11 | 0.9757 |
|  | 10% | -0.0089 | 0.4304 | 15.27 | 0.9867 |

Note: ECP (empirical coverage probability)

**TABLE 2.** Summary of Simulation Study Results: The table provides the posterior mean estimate, empirical bias, MSE, standard deviation, and coverage probability for 95% credible intervals, averaged over all 500 datasets.

| Parameter | % Reported | Estimate | Bias | MSE | SD | 95% ECP |
| --- | --- | --- | --- | --- | --- | --- |
| *Early Phase (March 6 to May 1), $T = 57$* |  |  |  |  |  |  |
| $\gamma_h$ | 100% | 0.1017 | 0.0017 | 0.0005 | 0.0229 | 0.9580 |
|  | 10% | 0.1009 | 0.0009 | 0.0005 | 0.0231 | 0.9500 |
| $\gamma_v$ | 100% | 0.4039 | 0.3039 | 0.1153 | 0.2486 | 0.7920 |
|  | 10% | 0.4103 | 0.3103 | 0.1163 | 0.2556 | 0.8160 |
| *Later Phase (March 6 to July 1), $T = 118$* |  |  |  |  |  |  |
| $\gamma_h$ | 100% | 0.1000 | 0.0000 | 0.0000 | 0.0052 | 0.9720 |
|  | 10% | 0.0995 | -0.0005 | 0.0000 | 0.0056 | 0.9760 |
| $\gamma_v$ | 100% | 0.1077 | 0.0077 | 0.0003 | 0.0192 | 0.9720 |
|  | 10% | 0.1093 | 0.0093 | 0.0004 | 0.0190 | 0.9620 |

Note: ECP (empirical coverage probability)

**FIGURE 2.**
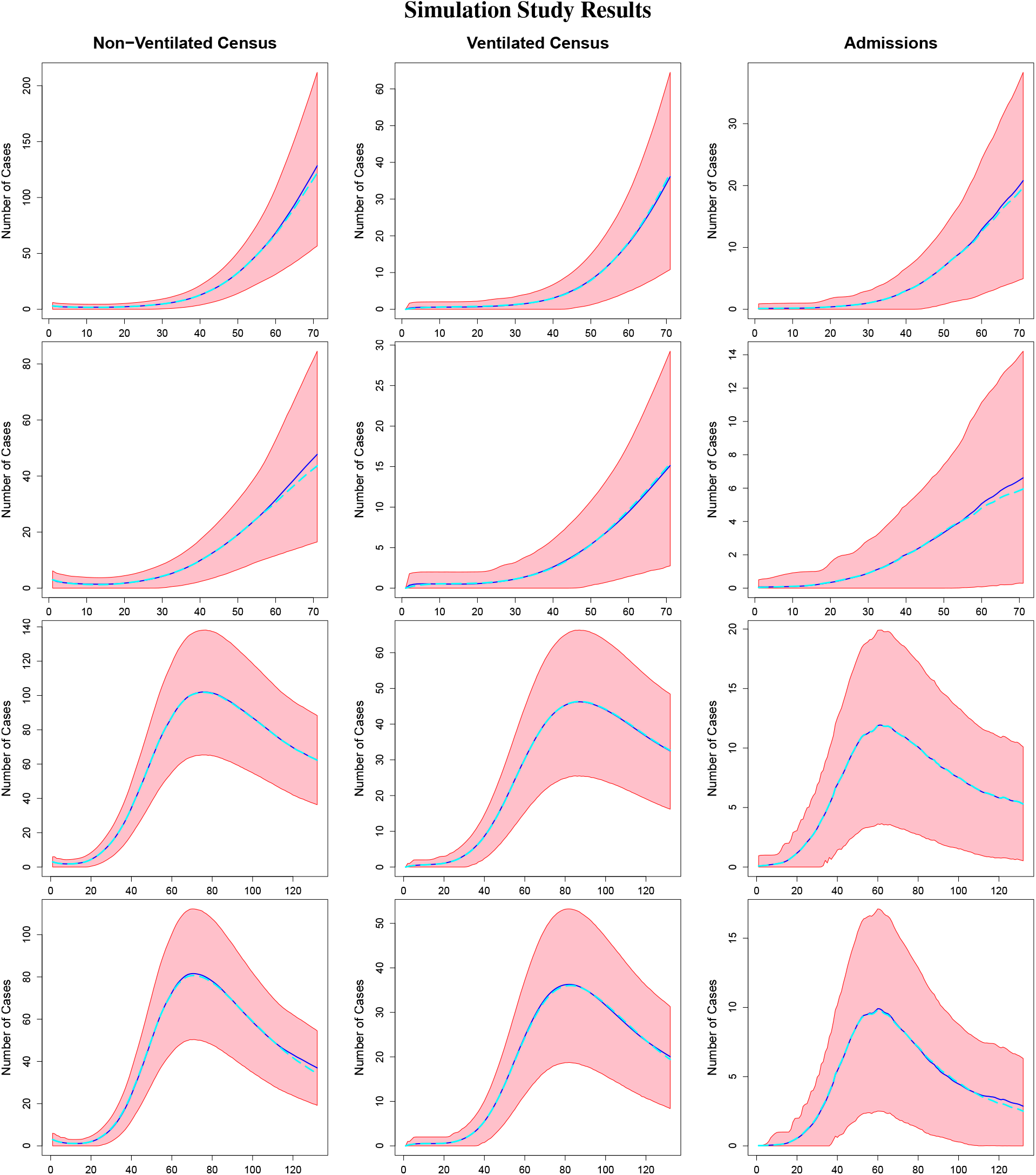
Simulation Study Results: The figure displays the posterior median (dark blue), true value used for data generation (light blue) and 95% prediction interval (red) for the non-ventilated census (column 1), ventilated census (column 2), and admissions (column 3) from the simulation with *T* = 57, and 100% detection (row 1), *T* = 57 and 10% detection (row 2), *T* = 118 and 100% detection (row 3) and *T* = 118 and 10% detection (row 4).

We find that our method is able to accurately predict the non-ventilated census, ventilated census, and number of SARS-CoV-2 positive admissions for the upcoming 14 day period with a high degree of accuracy, even in the presence of a large number of undetected cases. The bias in the point estimators for these quantities is relatively small, and mean absolute out of sample prediction error is less than 20% in most cases. As expected, the performance of the model (measured by the mean percent absolute out of sample prediction error) improves as the pandemic progresses and more data is available to the model. Our method is able to estimate the case recovery rate, the hospitalization recovery rate, and the ventilator recovery rate with small bias and MSE, even when there is under-detection.

Assuming full case detection, the methodology accurately estimates the area-level case incidence. As expected, in the presence of under-detection, there is significance bias in area-level incidence estimates (see Web Appendix C for these results). However, even when 100% of cases are detected, the empirical coverage intervals for the case recovery rate (*γ*_*i*_) and the area-level transmission rates (*β*_*c*_(*t*)*s*) are below their nominal levels, perhaps indicating a mild identifiabilty issue between these parameters. This is not surprising, given that an increase in the number of initially infected individuals can be offset by a decrease in the transmission rate or a increase in the recovery rate to produce the same number of incident infections (particularly during the early phases of a pandemic). Our primary goal is to predict the number of COVID-19 patients requiring inpatient care, not to perform inference with respect to model parameters. As the identifiability issue does not seem to impact the quantities of primary interest, it is of minimal concern for this application. We strongly caution the reader, however, against attempting to use this model to perform inference with respect to the recovery rate, transmission rate or the number of initially infected individuals.

## 4 PREDICTIVE ASSESSMENT

In this section, we assess the predictive performance of the Bayesian SIHVR model for the Prisma Health Upstate System and the Prisma Health Midlands System. Since the two regional systems serve geographically distinct patient populations, we fit separate models to the data from each region. The model from the Upstate system included census and admissions data summed over all hospitals in the Prisma Upstate system, and county-level incidence and social distancing data from Greenville and Spartanburg counties. While the Prisma Health Upstate system serves patients across the Upstate of South Carolina, Greenville and Spartanburg counties are the most populous. The model for the Midlands system included census and admissions data summed over all hospitals in the Midlands system and county-level incidence and social distancing data from Kershaw, Lexington, Richland and Sumter counties. To assess our model’s predictive performance during various pandemic stages, we fit the model using data from March 6th to May 1st, May 15th, June 1st, June 15th, and July 1st, for a total of 5 model fits per region. Each model was used to predict the daily reported case incidence, daily hospital admissions, daily non-ventilated census and daily ventilated census for the upcoming two week period. These predictions were compared to observed values to assess predictive performance. We also evaluated three different methods for incorporating social distancing metrics into the transmission rate. Method 1 took ***z***_*ct*_ as the scalar *z*_*c,t*−14_, the USDS social distancing metric from day *t* − 14. Method 2 took ***z***_*ct*_ = (*z*_*c,t*−14_, *z*_*c,t*−13_, …, *z*_*c,t*−3_)^′^, thereby allowing *β*_*c*_(*t*) to be influenced by the social distancing metric from days *t*−3 to *t*−14. Method 3 allowed for a more complex relationship between past levels of the social distancing metric and *β*(*t*). Specifically, method 3 took 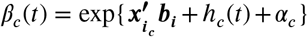, where 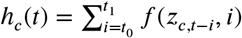, with *f* (*z,i*) being an unknown function that gives the influence of a social distancing level of *z* on day *t* − *i* on *β*(*t*). Note that *f* (*z,i*) is stationary with respect to *t*. The constants t0 and t1 are chosen reflect the range of past days whose social distancing metric affects *β*_*c*_(*t*), i.e., *β*_*c*_(*t*) is influenced by the amount of social distancing occurring between days *t* − *t*_1_ and *t* − *t*_0_, inclusive. The unknown function *f* (*z,i*) is approximated using the two dimensional tensor product of B-spline basis functions in the usual manner:

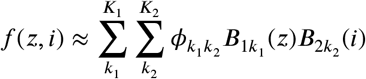

Yielding

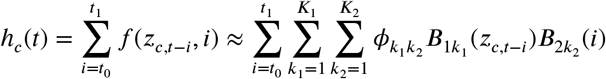

which implies

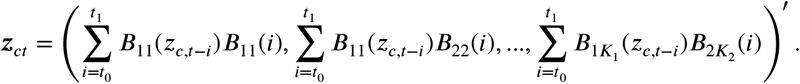

For our analysis, we took *t*_0_ = 3, *t*_1_ = 14, *K*_1_ = *K*_2_ = 3, and allowed the 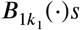 and 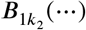 to be cubic B-spline basis functions with 3 equally spaced knots. Method 1 allows one to forecast up to two weeks into the future without predicting future levels of social distancing, an advantage over the more complicated methods. Creating such forecasts with method 2 or 3 requires first predicting future levels of social distancing; these future values were predicted via last value carried forward.

Table 3 summarizes the predictive performance of our model with respect to non-ventilated census, ventilated census, and daily SARS-CoV-2 positive admissions for each of the 3 methods for incorporating social distancing. The table provides the mean absolute prediction error for each of these quantities (defined as the absolute difference between the observed quantity and its predicted median, averaged over all 14 days for which predictions were computed). We also provide the percent of predicted days for which the 95% highest posterior density prediction interval for each quantity captures the observed value. The average number of days for which the observed non-ventilated census fell within the 95% prediction interval were 99.29%, 92.29%, and 100% for methods 1, 2, and 3 respectively. The average number of days for which the observed ventilated census fell within the 95% prediction interval were 84.29.%, 87.14%, and 86.64% for methods 1, 2 and 3, with the 95% prediction intervals for the number of admissions capturing the observed values 97.86%, 95.71%, and 95.00% on days on average for the three methods. While no single method consistently had the smallest prediction error across quantities and models, Method 1 had the lowest total prediction error (summed over all quantities and models). Figures 3 and 4 summarize the results from applying method 1 to the Upstate and Midlands data, respectively. Specifically, the figures provide the median estimate, 95% prediction interval, and observed data for the non-ventilated census, ventilated census, and number of admissions for each of the 5 time periods. Comparing the model predictions to the observed data for the 14 day forecast periods reveals that the model generally provides accurate predictions, with performance improving as more and more data is provided to the model. Importantly, the prediction intervals accurately quantify the uncertainty in model predictions. South Carolina experienced a rapid surge in COVID-19 cases in June and early July ^12^, with a corresponding in increase in COVID-19 hospitalizations. Our model successfully predicted this surge in both the Upstate and the Midlands region.

**TABLE 3.** The table provides the mean absolute prediction error and percent of observations falling with the 95% prediction interval (CP) for the non-ventilated census (NC), ventilated census (VC), and admissions (AD) for each of the 15 models fit to the data from the Upstate System (top) and Midlands system (bottom). Data from March 6th, 2020 to the day given in the Date column was used to fit each model, and the next 14 days of data was used to assess prediction.

| Date | Method | NC Pred. Er | NC CP | VC Pred. Er | VC CP | AD Pred. Er. | AD CP |
| --- | --- | --- | --- | --- | --- | --- | --- |
| *Upstate System* |  |  |  |  |  |  |  |
| May 1 | 1 | 8.29 | 1.00 | 1.07 | 1.00 | 4.93 | 1.00 |
|  | 2 | 3.14 | 1.00 | 0.64 | 1.00 | 2.93 | 1.00 |
|  | 3 | 15.00 | 1.00 | 1.36 | 1.00 | 8.00 | 0.86 |
| May 15 | 1.00 | 14.50 | 0.93 | 3.64 | 1.00 | 4.07 | 1.00 |
|  | 2 | 10.14 | 1.00 | 3.43 | 1.00 | 2.71 | 1.00 |
|  | 3 | 12.79 | 1.00 | 3.64 | 1.00 | 4.00 | 1.00 |
| June 1 | 1 | 4.86 | 1.00 | 7.79 | 0.29 | 3.07 | 0.93 |
|  | 2 | 8.36 | 1.00 | 8.07 | 0.29 | 3.71 | 0.86 |
|  | 3 | 4.36 | 1.00 | 7.79 | 0.29 | 3.57 | 0.93 |
| June 15 | 1.00 | 5.49 | 1.00 | 3.43 | 1.00 | 8.57 | 1.00 |
|  | 2 | 12.00 | 1.00 | 4.00 | 1.00 | 4.50 | 1.00 |
|  | 3 | 9.71 | 1.00 | 3.57 | 1.00 | 10.86 | 1.00 |
| July 1 | 1 | 7.93 | 1.00 | 7.64 | 0.86 | 5.14 | 1.00 |
|  | 2 | 42.50 | 1.00 | 6.07 | 1.00 | 19.14 | 1.00 |
|  | 3 | 18.64 | 1.00 | 4.43 | 1.00 | 12.14 | 1.00 |
| *Midlands System* |  |  |  |  |  |  |  |
| May 1 | 1.00 | 5.71 | 1 | 11.71 | 0.76 | 3.36 | 1.00 |
|  | 2 | 25.29 | 0.29 | 4.5 | 1.00 | 2.71 | 0.79 |
|  | 3 | 15.00 | 1.00 | 8.57 | 1.00 | 1.86 | 1.00 |
| May 15 | 1 | 18.14 | 1.00 | 17.36 | 0.50 | 6.50 | 0.93 |
|  | 2 | 22.86 | 1.00 | 18.93 | 0.43 | 7.64 | 1.00 |
|  | 3 | 24.43 | 1.00 | 19.29 | 0.50 | 7.93 | 0.86 |
| June 1 | 1 | 8.79 | 1.00 | 1.71 | 1.00 | 2.71 | 0.93 |
|  | 2 | 9.43 | 1.00 | 1.64 | 1.00 | 2.79 | 0.93 |
|  | 3 | 11.93 | 1.00 | 1.79 | 1.00 | 3.29 | 0.86 |
| June 15 | 1 | 4.21 | 1.00 | 3.14 | 1.00 | 2.64 | 1.00 |
|  | 2 | 3.71 | 1.00 | 2.64 | 1.00 | 2.86 | 1.00 |
|  | 3 | 5.07 | 1.00 | 3.21 | 1.00 | 2.57 | 1.00 |
| July 1 | 1 | 7.41 | 1.00 | 5.42 | 1.00 | 2.78 | 1.00 |
|  | 2 | 8.50 | 1.00 | 5.43 | 1.00 | 3.00 | 1.00 |
|  | 3 | 8.14 | 1.00 | 5.21 | 1.00 | 2.79 | 1.00 |

**FIGURE 3.**
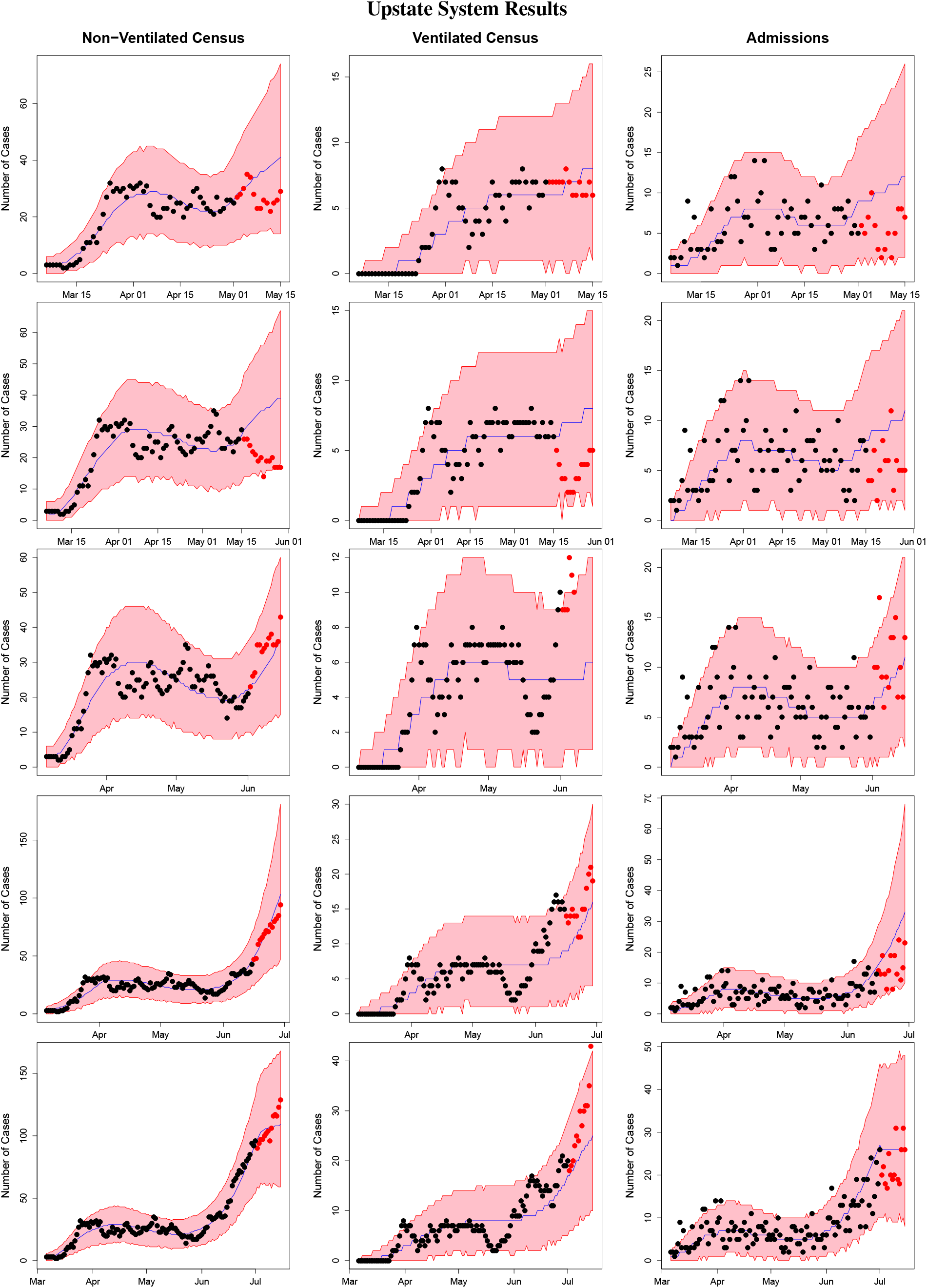
The figure displays the results for the non-ventilated census (column 1), ventilated census (column 2), and SARS-CoV-2 positive admissions (column 3) for the Upstate system from the models fit using data from March 6th to May 1st (row 1), May 15th (row 2), June 1 (row 3), June 15 (row 4), and July 1 (row 5). The red shaded regions denote 95% prediction intervals, the blue lines denote the median estimators, the black points denote the observed data used to fit the model, and the red points denote observed data from the 14 forecast period (not used to fit the model).

**FIGURE 4.**
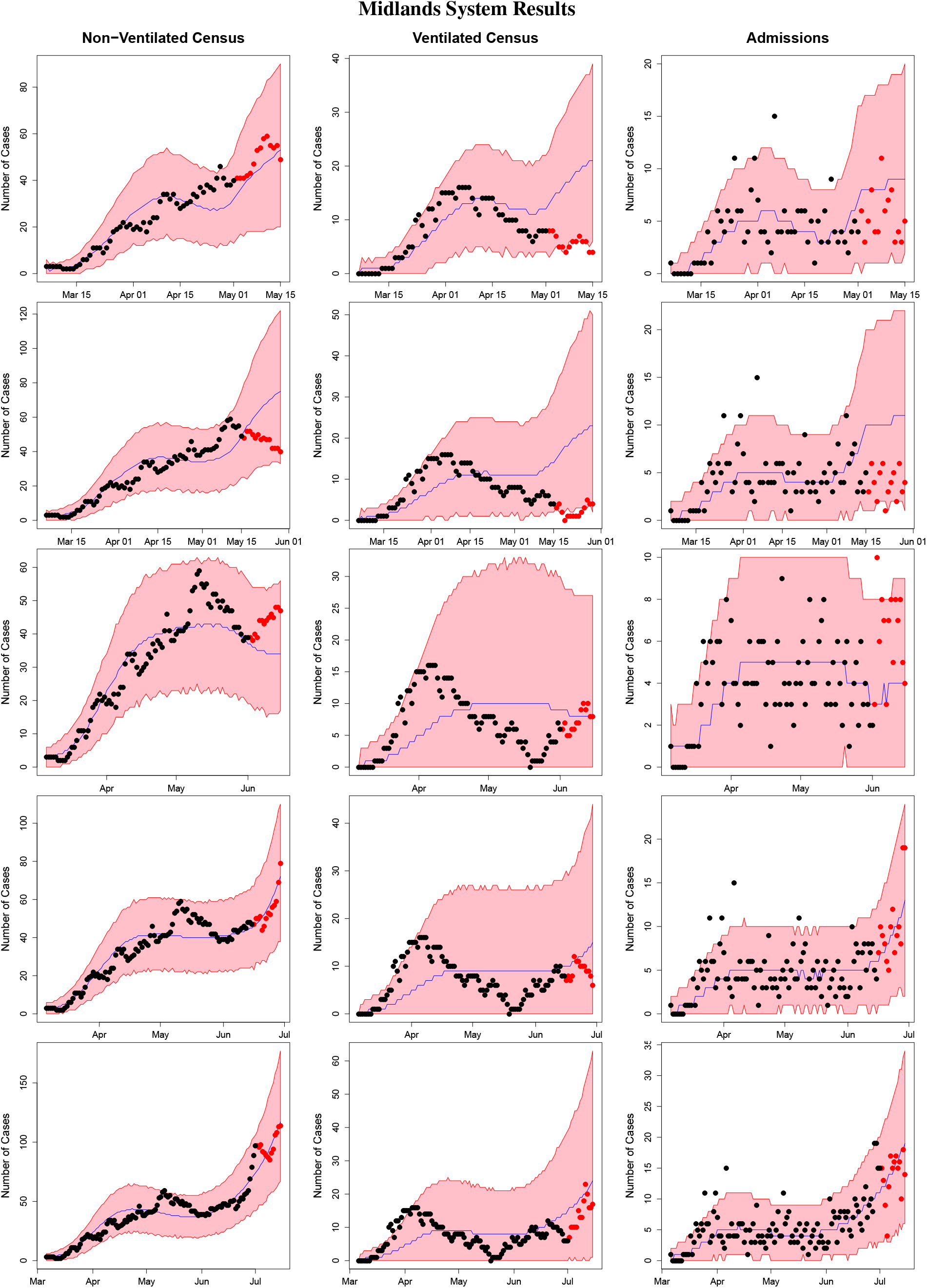
The figure displays the results for the non-ventilated census (column 1), ventilated census (column 2), and SARS-CoV-2 positive admissions (column 3) for the Midlands system from the models fit using data from March 6th to May 1st (row 1), May 15th (row 2), June 1 (row 3), June 15 (row 4), and July 1 (row 5). The red shaded regions denote 95% prediction interval, the blue lines denote the median estimators, the black points denote the observed data used to fit the model, and the red points denote observed data from the 14 forecast period (not used to fit the model).

Web Figures 3 and 4 in Web Appendix C provide the estimates of the number of daily COVID-19 cases reported by each county in Upstate and Midlands System from each model fit (using method 1).

### 4.1 Performance Comparison

To further assess the performance of our methodology, we compared its performance to that of the Institute for Health Metrics and Evaluation (IHME) model. The IHME model provides national and state level estimates of daily COVID-19 deaths, COVID-19 cases, inpatient beds occupied by COVID-19 patients, and ventilators in use by COVID-19 patients. As reliable data on the number of SARS-CoV-2 positive patients admitted to inpatient care each day is not readily available for the entire state of South Carolina, and reliable data on the number of ventilators in use by COVID-19 patients in South Carolina is not available for the early months of the pandemic, we elected to compare the following simplified version of our model to the IHME model’s prediction for South Carolina. We assumed only the statewide case incidence and statewide hospital census data were observed:

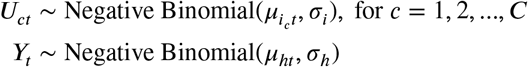

and solved simpler system of differential equations without the ventilated state:

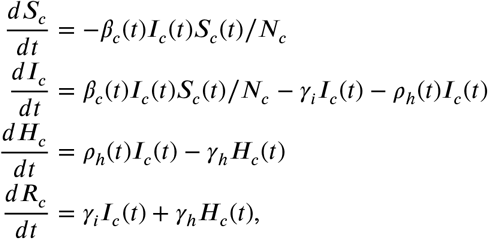

which we refer to as the Susceptible-Infectious-Hospitalized-Recovered (SIHR) model. The functional forms of *β*_*c*_(·) and *ρ*_*h*_(·) and the prior distributions were identical to those described in Section 2, and method 1 was used to incorporate the social distancing data. However, here *C* = 1, and *U*_*ct*_ and *Y*_*t*_ are the number of reported COVID-19 cases and hospital beds occupied by COVID-19 patients on day *t* for the entire state of South Carolina, as reported by SCDHEC ^12^. The social distancing metric was Unacast’s ‘Change in Visits to Non-Essential Businesses’ metric for the entire state of South Carolina. We fit our model using data from March 6th, to July 1st, predicted out 2 weeks, and compared the results to the IHME’s reference model projections published on July 7th. Figure 5 displays the number hospital beds occupied and the number of reported infections, along with the corresponding predictions from our model and IHME’s model. As can clearly be seen, the Bayesian SIHVR model provides more accurate predictions for both quantities. The IHME model has several strengths, including the fact that it produces estimates for every US state and many other countries using only data which is readily and widely available. This gives policy makers a way to easily compare predictions at different locations and allocate resources accordingly. However, the performance of our method shows that incorporating more (local) data sources can provide more accurate, locally tailored predictions for individual states or healthcare systems. Importantly, our method more accurately predicted the rapid surge in COVID-19 cases which took place in South Carolina during June and early July of 2020, while the IHME model significantly underestimated the number of individuals requiring hospitalization during this surge.

**FIGURE 5.**
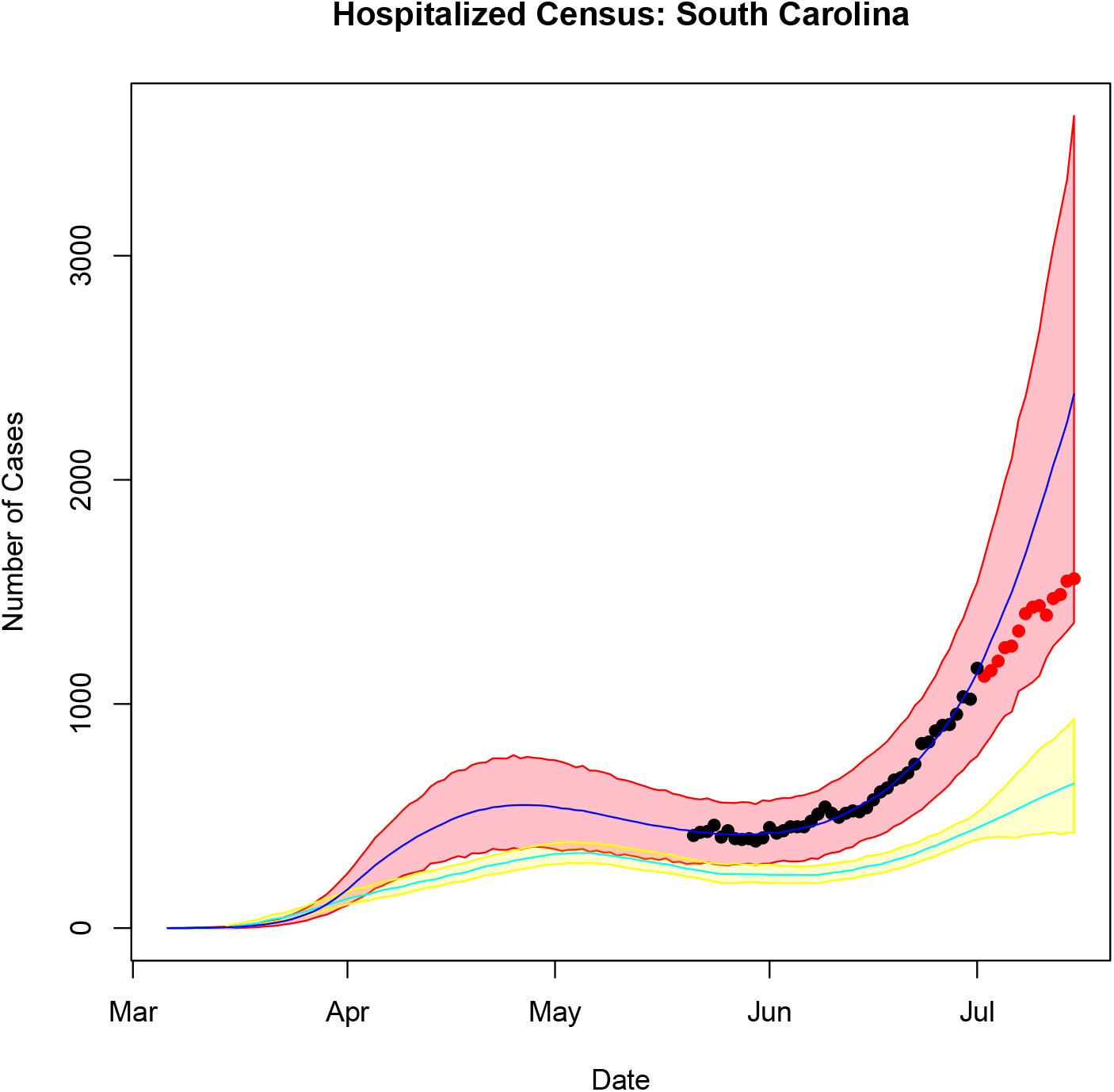
The figure compares the performance of the Bayesian SIHR model to the that of the IHME model. Depicted are the median (dark blue) and 95% prediction interval (red) from the Bayesian SIHR model and the mean (light blue) and 95% prediction interval (yellow) from the IHME model for the hospitalized census. Data points used to fit the Bayesian SIHR model are shown in black, and data points withheld to assess predictive performance are shown in red.

## 5 CONCLUSION

As the United States continues to ease social distancing restrictions, the need to predict the burden of COVID-19 on local healthcare systems is likely to persist. The ongoing return to in-person instruction in many schools and universities, along with the possibility of increased viral transmission in the colder winter months may lead to continued COVID-19 resurgence in the US ^19,20^. Thus far, the severity of the COVID-19 pandemic has displayed substantial heterogeneity among states, with Northeastern states such as New York and New Jersey being the hardest hit in the early stages of the pandemic, with Southern and Southwestern states such as Florida, South Carolina, Texas and Arizona seeing larger outbreaks during the summer months. This heterogeneity makes it essential to produce reliable, locally customizable forecasts of COVID-19 burden so that resources can be transferred to where they are needed.

We have presented a method for forecasting the demand for inpatient COVID-19 care at the healthcare system level which is locally customizable. Our Bayesian SIHVR model incorporates local COVID-19 reported case incidence, local social distancing patterns, and healthcare system-specific COVID-19 data. Our model provides accurate short-term (2 week) predictions and reliable regions of uncertainty. To facilitate the further use of our model by healthcare systems, code which fits the model has been developed in R and made available on GitHub (https://github.com/scwatson812/BayesianSIHVRModel).

## Data Availability

The USDS data is publicly available on the Unacast webpage at https://www.unacast.com/covid19/social-distancing-scoreboard. The county and state level reported case incidence data, and statewide hospital census data is publicly available on the SCDHEC webpage at https://scdhec.gov/covid19/south-carolina-county-level-data-covid-19.
Code which implements the model described in the manuscript may be found at https://github.com/scwatson812/BayesianSIHVRModel

## ACKNOWLEDGMENTS

The authors would like to thank the Prisma Health System for their support for this project and for providing the data.

## Author contributions

SS and AM developed the methods. RH, SA, and JE collected and cleaned the data. SS and RH fit the models. SS, CR, and AM contributed to writing the manuscript. All authors reviewed the manuscript.

## Data Accessibility

The USDS data is publicly available on the Unacast webpage at https://www.unacast.com/covid19/social-distancing-scoreboard. The county and state level reported case incidence data, and statewide hospital census data is publicly available on the SCD-HEC webpage at https://scdhec.gov/covid19/south-carolina-county-level-data-covid-19. For easier extraction, this data has been formatted as a csv file and made available in the journal’s data repository.

## SUPPORTING INFORMATION

The following supporting information is available as part of the online article: a description of the MCMC sampling algorithm (Web Appendix A), additional details regarding the simulation study (Web Appendix B), and additional details and figures regarding estimation of reported area-level COVID-19 case incidence with the Bayesian SIHVR model (Web Appendix C).

