## Supplemental Material for "A Bayesian Susceptible-Infectious-Hospitalized-Ventilated-Recovered Model to Predict Demand for COVID-19 Inpatient Care in a Large Healthcare System"

#### 1 Web Appendix A: Markov Chain Monte Carlo Algorithm for Posterior Sampling

Details of the Markov chain Monte Carlo algorithm used to sample from the posterior distribution of the Bayesian SIHVR model are provided in Algorithm 1. All parameters are sampled from their full conditional distributions with Metropolis Hastings steps. Evaluating the full conditional distribution each parameter requires solving the SIHVR system of differential equations using the most recently sampled values of all unknown parameters. The Euler method is used to provide a computationally efficient way to repeatedly solve the system of differential equations.

**ALGORITHM 1:** The Markov chain Monte Carlo algorithm used to fit the Bayesian SIHVR model. Here $P_i$, $P_d$, $P_h$, and $P_v$ denote the dimensions of $\boldsymbol{b}_i$, $\boldsymbol{d}$, $\boldsymbol{b}_h$ and $\boldsymbol{b}_v$, respectively.

Initialize $\boldsymbol{\theta}^{(0)}$ for all unknown parameters $\boldsymbol{\theta}$;
**for** g $\leftarrow$ 1 **to** $G$
  **for** $j \leftarrow 1$ **to** $\mathrm{P}_i$
    Sample ${b_i}_j^{(g)}$;
  **for** j $\leftarrow$ 1 **to** $\mathrm{P}_d$
    Sample $d_j^{(g)}$;
  Sample $\gamma_i^{(g)}$;
  Sample $\gamma_h^{(g)}$;
  Sample $\gamma_v^{(g)}$;
  **for** c $\leftarrow$ 1 **to** C
    Sample $\alpha_c^{(g)}$;
  **for** c $\leftarrow$ 1 **to** C
    Sample $\mathrm{I}_c(0)^{(g)}$;
  **for** j $\leftarrow$ 1 **to** $\mathrm{P}_h$
    Sample ${b_h}_j^{(g)}$;
  **for** j $\leftarrow$ 1 **to** $\mathrm{P}_v$
    Sample ${b_v}_j^{(g)}$;
  Sample $\sigma_i^{(g)}$;
  Sample $\sigma_a^{(g)}$;
  Sample $\sigma_h^{(g)}$;
  Sample $\sigma_v^{(g)}$;
  **if** $g$ mod $100 = 0$
  **then** Tune the proposal distributions;

#### 2 Web Appendix B: Additional Details Regarding the Simulation Study

This section provides additional details regarding the parameter specifications used for data generation in the simulation study. For each for the four simulation configurations, the parameters were chosen so that generated data resembled the data observed during the Upstate system in the early phase (March 6th-May 15st) for $T = 57$ or later phase (March 6th-July 1st) for $T = 118$.

For all configurations, we assumed there was one initially infectious non-hospitalized individual in each county, 1.5 hospitalized individuals from each county (chosen to sum the total of 3 hospitalized individuals in the Upstate system on

March 5th), 0 ventilated individuals in each county (chosen to sum the total of 0 ventilated individuals in the Upstate system on March 5th), and 0 recovered individuals in each county. The remaining individuals were assumed to be susceptible, giving the following initial conditions: $S_1(0) = 498399.5$, $S_2(0) = 302192.5$, $I_1(0) = I_2(0) = 1$, $H_1(0) = H_2(0) = 1.5$, $V_1(0) = V_2(0) = R_1(0) = R_2(0) = 0$. The county level random effects were taken to be $\alpha_1 = 0$, $\alpha_2 = 0.1$. The proportion of hospitalized patients entering the ventilated state each day was taken to be 0.05 (i.e. $\rho_v(t) = 0.05$). Web Figure 1 shows the values of $\rho_h(t)$ used for each simulation configuration, and Web Figure 3 shows the values of $\beta_c(t)$, $c = 1, 2$, used for each simulation configuration.

### 3 Web Appendix C: Additional Details Regarding Estimation of Reported COVID-19 Cases

This section provides additional details regarding estimation of area-level reported case incidence using the Bayesian SIHVR model. While such estimation was not our primary goal, it may be of interest for healthcare systems to understand the local trajectory of the pandemic in their area. Tables 1 and 2 and Figure 2 summarize the results of the simulation study described in Section 3 of the manuscript with regard to reported case incidence, and the associated transmission rate ($\beta_c(t)$) and recovery rate $\gamma_i$. Specifically, Table 1 provides the empirical bias, absolute prediction error, percent absolute prediction error, and 95% empirical coverage probability (ECP) for the reported case incidence from each area. The empirical bias is averaged over days $t = 1, 2, ...T$, and the other quantities are averaged over days $t = T+1, T+2, ..., T+14$. As the posterior distribution of the $\mu_{i_c,t}$s was right skewed, the posterior median was used as a point estimate. Empirical bias, absolute prediction error, and percent absolute prediction error were calculated with respect to the true case incidence, that is, with respect to the values of $\mu_{i_c,t}$ obtained from solving the SIHVR system of differential equations. However, 95% ECP was calculated using the *reported* case incidence, i.e., for $c = 1, 2$ and $t = T+1, ..., T+14$, $U_{ct}$ was generated from a Poisson distribution with mean $\mu_{i_c,t} * r$, where $r = 1$ under the assumption of 100% case detection and $r = 0.1$ under the assumption of 10% case detection; ECPs were then calculated by assessing the how often the generated $U_{ct}$ value fell within the corresponding 95% prediction interval. Table 2 provides the posterior mean estimate, empirical bias, MSE, standard deviation, and 95% ECP for the recovery rate $\gamma_i$. Figure 2 provides the posterior point estimate, true parameter value, and 95% prediction interval (averaged over all 500 datasets) for the reported case incidence and transmission rate for each area in the simulation. The point estimator for the case incidence and transmission rate are the posterior median and mean, respectively.

Our simulation study found that the Bayesian SIHVR model was able to accurately estimate the daily number of new reported COVID-19 cases. Under the assumption of complete case detection, the model also accurately estimates the true case incidence. However, even in the presence of under-detection, the model accurately predicts *reported* case incidence (as evidenced by the empirical coverage for 95% credible prediction intervals for reported case incidence). In the presence of under-detection, the number of confirmed cases is a severely biased estimator of the true case numbers, and appears to underestimate them by a factor of 10, as one would expect given that only 10% of cases are detected.

Figures 3 and 4 display the county-level reported case incidence results from applying the Bayesian SIHVR model (method 1) to data from the Upstate and Midlands Systems, respectively. The figures provide the posterior median (blue), reported case incidence used to fit the model (black), reported case incidence withheld to assess predictive performance (red), and 95% prediction intervals (shaded red). It is notable that the Bayesian SIHVR model accurately predicted the surge in reported cases which took place in June and early July for most counties.

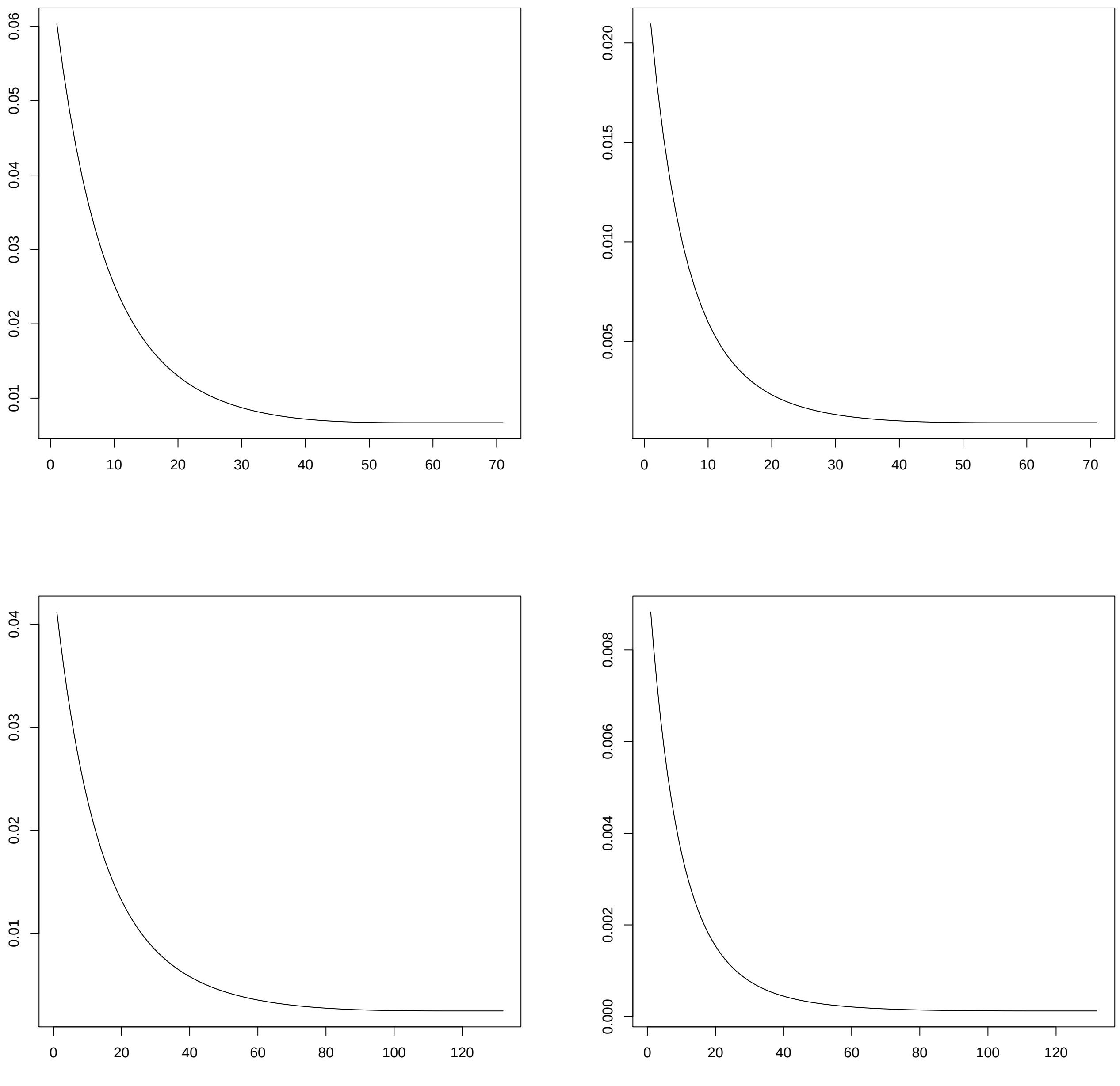


Web Figure 1: The figure displays the proportion of individuals in the infectious state entering the hospitalization state each data ($\rho_h(t)$) used for data generation in the simulation configuration with $T = 57$ (top row) and $T = 118$ (bottom row). The columns correspond to 100% detection (left) and 10% detection (right).

| Quantity | % Reported | Bias | Abs. Pred. Er. | % Abs. Pred. Er. | 95% ECP |
|---|---|---|---|---|---|
| | | *Early Phase (March 6 to May 1), T = 57* | | | |
| County 1 | 100% | 0.0027 | 9.7687 | 8.57 | 0.9751 |
| | 10% | -66.4626 | 193.4928 | 89.63 | 0.9644 |
| County 2 | 100% | -0.0419 | 14.5346 | 8.34 | 0.9779 |
| | 10% | -98.5124 | 295.8364 | 89.42 | 0.9636 |
| | | *Later Phase (March 6 to July 1), T = 118* | | | |
| County 1 | 100% | 0.0159 | 2.1869 | 4.28 | 0.9667 |
| | 10% | -903.9671 | 722.7815 | 90.52 | 0.9371 |
| County 2 | 100% | 0.0363 | 3.9706 | 4.10 | 0.9733 |
| | 10% | -987.0781 | 398.2774 | 87.21 | 0.7871 |

Web Table 1: Summary of Simulation Study Results: The table provides the empirical bias (averaged over days $1, 2, ..., T$ and the 500 datasets), empirical mean absolute prediction error (averaged over days $T + 1, T + 2, ..., T + 14$ and the 500 datasets), empirical mean percent absolute prediction error (averaged over averaged over days $T + 1, T + 2, ..., T + 14$ and the 500 datasets), and empirical coverage probability for 95% forecast prediction intervals (averaged averaged over days $T + 1, T + 2, ..., T + 14$ and the 500 datasets) for the reported case area-level case incidence.

| Parameter | % Reported | Estimate | Bias | MSE | SD | 95% ECP |
|---|---|---|---|---|---|---|
| | | *Early Phase (March 6 to May 1), T = 57* | | | | |
| $\gamma_i$ | 100% | 0.0645 | -0.0070 | 0.0006 | 0.0094 | 0.4640 |
| | 10% | 0.0540 | -0.0174 | 0.0014 | 0.0135 | 0.4140 |
| | | *Later Phase (March 6 to July 1), T = 118* | | | | |
| $\gamma_i$ | 100% | 0.0710 | -0.0004 | 0.0000 | 0.0022 | 0.5360 |
| | 10% | 0.0743 | 0.0029 | 0.0000 | 0.0023 | 0.4300 |

Web Table 2: Summary of Simulation Study Results: The table provides the empirical bias (averaged over days $1, 2, ..., T$ and the 500 datasets), empirical mean absolute prediction error (averaged over days $T + 1, T + 2, ..., T + 14$ and the 500 datasets), empirical mean percent absolute prediction error (averaged over averaged over days $T + 1, T + 2, ..., T + 14$ and the 500 datasets), and empirical coverage probability for 95% forecast prediction intervals (averaged averaged over days $T + 1, T + 2, ..., T + 14$ and the 500 datasets) for the reported case area-level case incidence.

#### Simulation Study Results

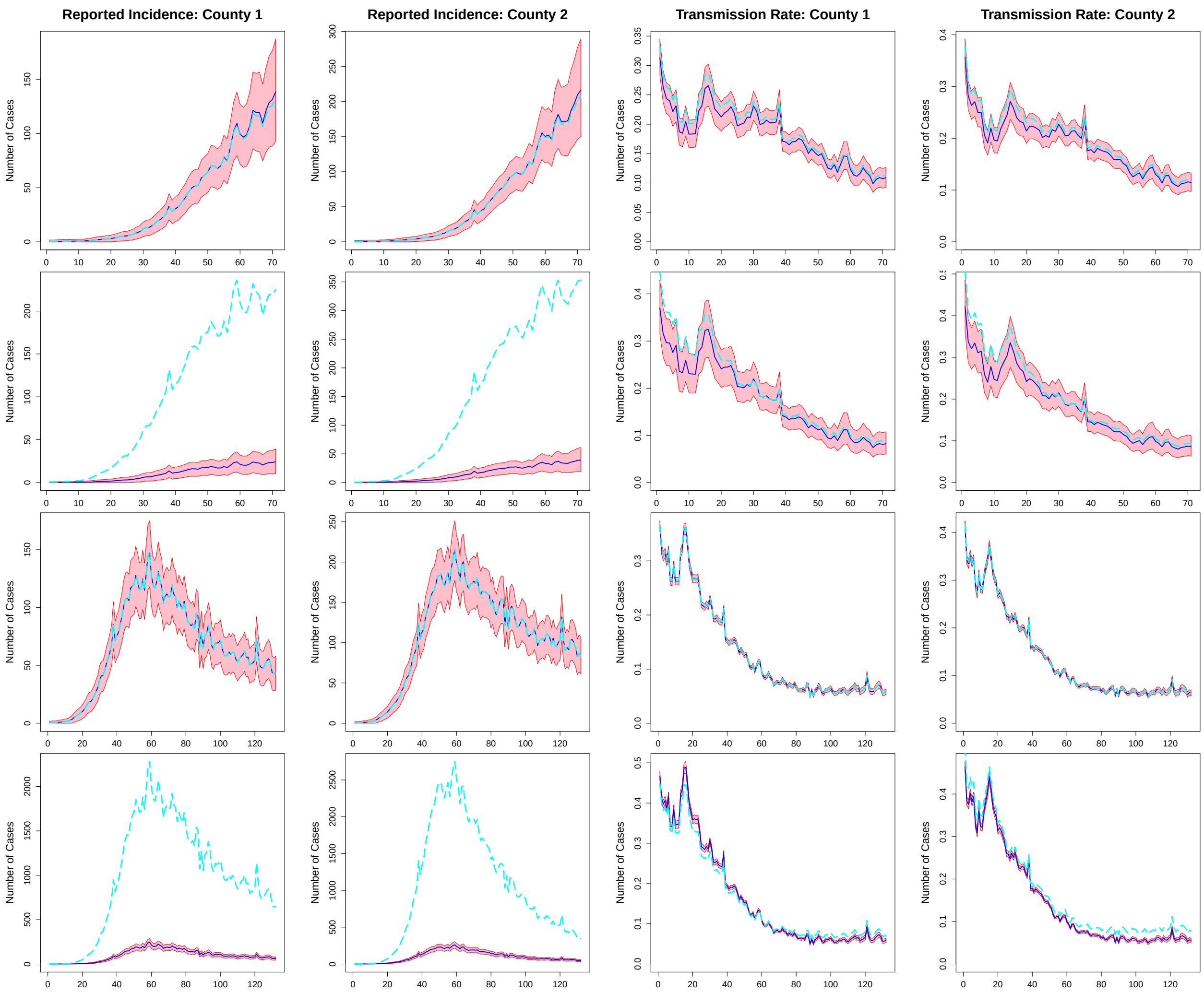


Web Figure 2: Simulation Study Results: The figure displays the posterior point estimate (median for reported incidence, mean for transmission rate, dark blue), true value used for data generation (light blue) and 95% credible interval (red) for the reported incidence in county 1 (column 1), reported incidence in county 2 (column 2), transmission rate in county 1 ($\beta_1(\cdot)$, column 3) and transmission rate in county 2 ($\beta_2(\cdot)$, column 4) from the simulation with $T = 57$, and 100% detection (row 1), $T = 57$ and 10% detection (row 2), $T = 118$ and 100% detection (row 3) and $T = 118$ and 10% detection (row 4). In the presence of under-detection, the light blue line on the county incidence plots is the true number of cases, not the reported number.

#### Upstate System County Results

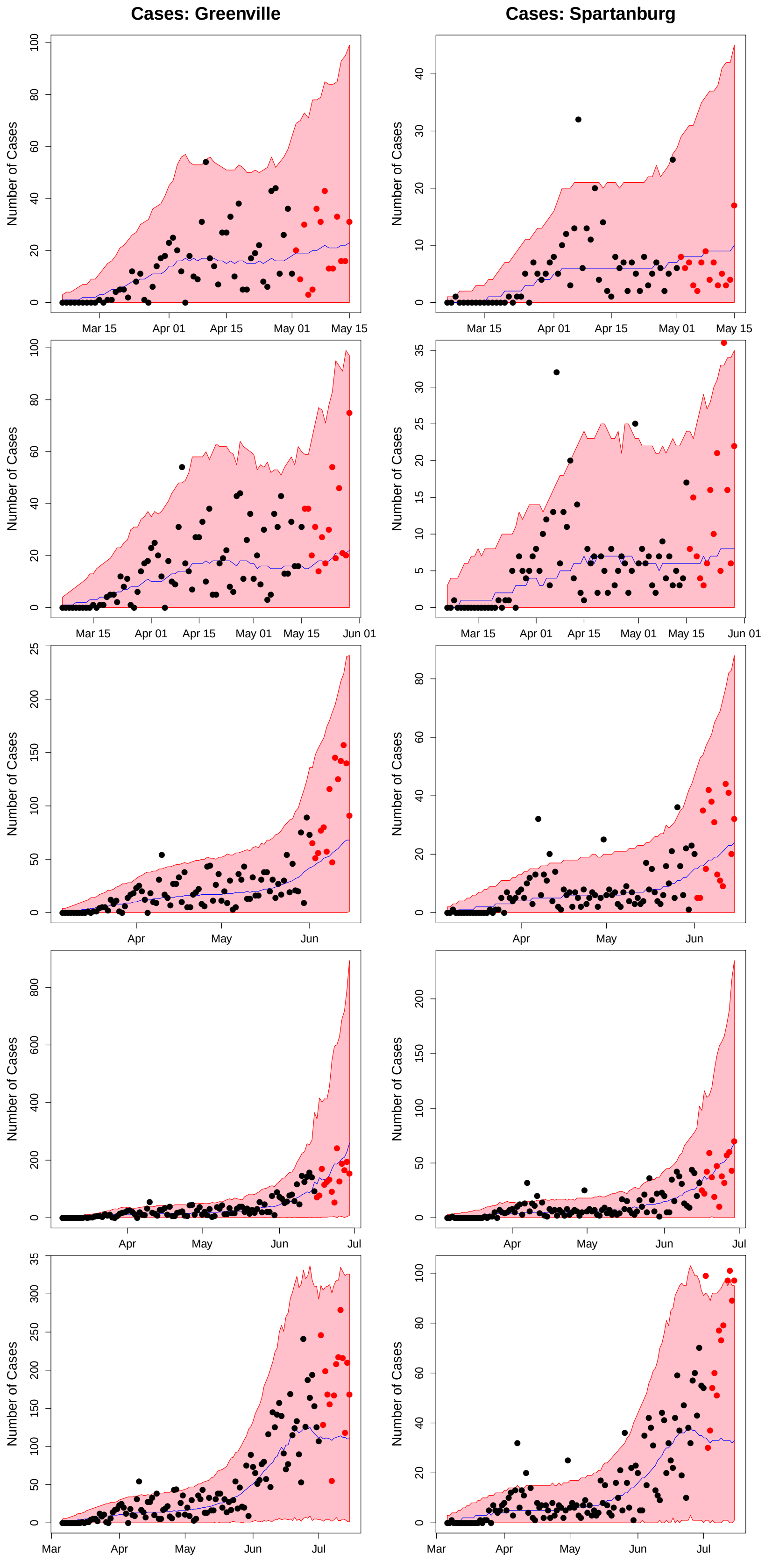


Web Figure 3: The figure displays the model predicted number of reported COVID-19 cases for the counties in the Prisma Health Upstate System from the models fit using data from March 6th to May 1st (row 1), May 15th (row 2), June 1st (row 3), June 15th (row 4) and July 1st (row 5). The red shaded regions denote 95% prediction intervals, the blue lines denote the median estimators, the black points denote the observed data used to fit the model, and the red points denote observed data from the 14 forecast period (not used to fit the model).

#### Midlands System County Results

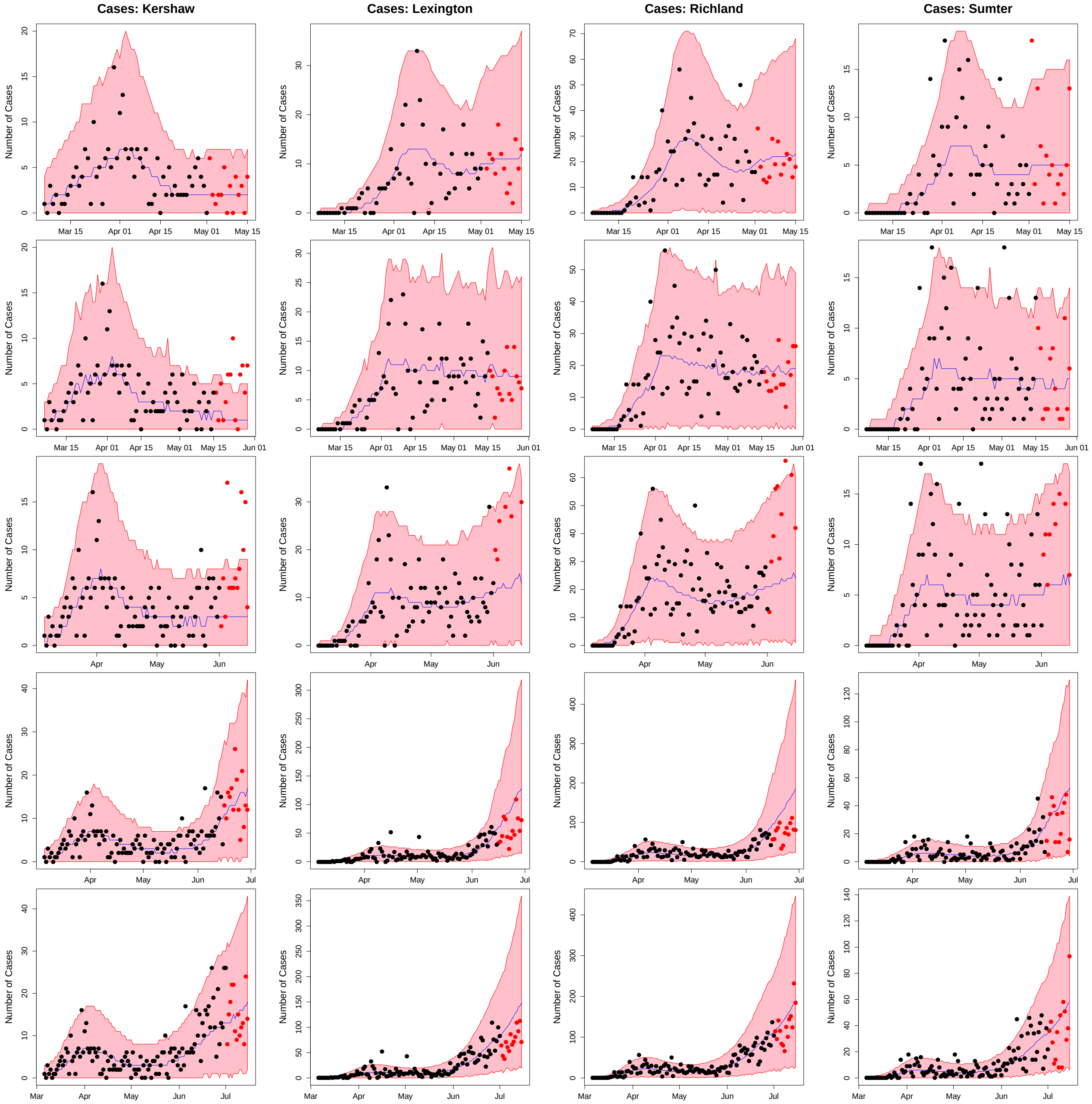


Web Figure 4: The figure displays the model predicted number of reported COVID-19 cases for the counties in the Prisma Health Midlands System from the models fit using data from March 6th to May 1st (row 1), May 15th (row 2), June 1st (row 3), June 15th (row 4) and July 1st (row 5). The red shaded regions denote 95% prediction intervals, the blue lines denote the median estimators, the black points denote the observed data used to fit the model, and the red points denote observed data from the 14 forecast period (not used to fit the model).
